# Integrating virtual reality, electroencephalography, and transcranial magnetic stimulation to study the neural origin of the sublime: The SUBRAIN protocol

**DOI:** 10.1101/2024.04.14.24305786

**Authors:** Elena Bondi, Flavia Carbone, Marta Pizzolante, Giandomenico Schiena, Adele Ferro, Maddalena Mazzocut-Mis, Andrea Gaggioli, Alice Chirico, Paolo Brambilla, Eleonora Maggioni

## Abstract

**Introduction:** Awe is a complex emotion unveiling a positive and mixed nature, which resembles the Romantic feeling of the Sublime. It has increasingly become the object of scientific investigation in the last twenty years. However, its underlying brain mechanisms are still unclear. To fully capture its nature in the lab, researchers have increasingly relied on virtual reality (VR) as an emotion-elicitation method, which can resemble even complex phenomena in a limited space. In this work, a multidisciplinary team proposed a novel experimental protocol integrating VR, electroencephalography (EEG), and transcranial magnetic stimulation (TMS) to investigate the brain mechanisms of this emotion.

**Methods:** A group of bioengineers, psychologists, psychiatrists, and philosophers designed the SUBRAIN study, a single-center, one-harm, non-randomized interventional study to explore the neural processes underlying awe experiences. The study will be performed on fifty adults. The experimental protocol includes different steps: (i) screening, (ii) enrollment, (iii) pre-experimental assessment, (iv) VR experimental assessment, and (v) post-experimental debriefing. The brain’s electrical activity is recorded using the EEG while participants navigated three immersive awe-inducing VR environments (VREs) and a neutral one. At the same time, the cortical excitability and connectivity is investigated by performing a TMS-EEG session right after each VR navigation. Along with cerebral signals, self-reported questionnaires were used to assess the VR-induced changes in the emotional state of the subjects. This data is then analyzed to delve into the cerebral mechanisms of awe.

**Discussion:** This study protocol is the first one that tries to fully understand the neural bases of awe by eliciting and studying this phenomenon in VR. The pairing of awe-inducing VR experiences and questionnaires investigating participants’ affect and emotions, with non-invasive neural techniques, can provide a novel and extensive knowledge on this complex phenomenon. The protocol can inform on the combination of different instruments showing a reproducible and reliable setting for the investigation of induced complex emotions.

## Introduction

The sublime – the mixed feeling of pleasure and displeasure – has its roots at the heart of the eighteenth century, in a precise philosophical tradition that leads to Kant’s theory, according to which the sublime has a systematic-transcendental nature and is a uniquely subjective feeling (1–3). This feeling-emotion is a combination of fear, enthusiasm, astonishment, wonder (4). Romanticism, furthermore, accentuates profound emotion as an authentic wellspring of aesthetic encounter (5). It bestows fresh significance upon occurrences of wonder and awe by legitimizing such sentiments as reactions to the sublime in nature.

In 2010, Richardson (6) establishes a connection between the eighteenth-century sublime (particularly that elaborated by Edmond Burke (1)) and neuroscience, while the following year, Onians (7) asserts that Burke did not know, as we do now, that the profound awe of Burkean origin occurs due to the operation of natural selection on our neural apparatus. In 2014, Ishizu and Seki report the results of a study conducted on 21 subjects to whom images of natural beauty and natural sublimity were shown while their brains were examined using fMRI technology (8). Ishizu and Seki’s researches demonstrates that natural beauty and natural sublimity activate different brain areas and evoke very different experiences in humans (8–10).

Recently, the sublime has increasingly become the subject of multidisciplinary and interdisciplinary investigations, combining philosophical and psychological approaches. A particular focus of psychology has been on awe as the subjective response to sublime stimuli (11,12), also besides the aesthetic domain (13,14). Despite initial neural evidence on the sublime stimuli (8), the understanding of awe’s underlying brain mechanisms remains at an early stage. Alike the sublime (15), awe emerged as an intense emotional state combining positive and negative components simultaneously (11,16), which arises in response to stimuli perceived as perceptually (e.g., waterfalls, trees, mountains, and tempests) (17) or conceptually (18) vast or overwhelming. Most studies have used awe-inducing videos or images to elicit awe in the lab, but these tend to elicit more subtle emotions with respect to those evoked by real-life experiences. In this context, virtual reality (VR) technology can be used to create a more intense laboratory version of awe (14,19). VR is capable of creating realistic scenarios (20) by also providing a multisensory stimulation. Compared to other emotion-inducing tools, VR provides a unique sense of presence and immersion and a heightened emotional intensity of the experience (21). In 2018, our research team already validated three awe-inducing immersive VR environments representing natural scenarios (i.e., forest, mountains, and earth view from deep space) (22), since nature is recognized as the most frequent trigger of awe-inspiring experiences (23), as compared to a non-awe-inducing natural environment with green grass, trees, and flowers.

Although the mechanisms underpinning awe experiences remain largely elusive, awe-inducing VR scenarios designed for a laboratory setting can allow us to identify experiential, behavioral, cognitive, and brain correlates of awe. So far, few studies have investigated the brain activity correlates of the awe emotion through functional Magnetic Resonance Imaging (fMRI), suggesting the involvement of the default mode network (DMN) within awe experiences (24) and showing a proportional activation of cortical (temporal and frontal) and subcortical (basal ganglia and cerebellum) brain areas to the sublime intensity (25). Moreover, a recent fMRI study suggested the left middle temporal gyrus as pivotal in processing both positive and threatening awe experiences and explored its interactions with other brain regions usually involved in emotional processing (26). Despite this preliminary fMRI evidence, a more profound comprehension of the neural underpinnings of the awe experience is needed. In this context, a deeper knowledge of the brain activity and connectivity patterns underlying this complex experience can be obtained by combining electroencephalography (EEG) and transcranial magnetic stimulation (TMS). TMS and EEG are non-invasive techniques that provide direct insights into brain networks’ instantaneous and directional interactions (27–30). Unlike fMRI, the EEG technique provides direct information on the brain’s electrical activity (31), boasting exceptional time resolution to explore functional brain connectivity on the neuronal time scale across multiple frequency bands (27,30), while TMS enhances the EEG potential for studying the propagation of information within the brain networks (28,29). By leveraging EEG to track TMS pulses through functionally linked brain regions, TMS-EEG integration emerges as a valuable tool to investigate cortical excitability and effective (directional) connectivity. Hence, TMS-EEG integration holds promise in unveiling how the propagation of neural information within the brain networks is affected by sublime emotional experiences elicited by means of nature-based VR scenarios.

Within this context, for the first time, the multidisciplinary SUBRAIN project investigates the psychological and neural correlates of different laboratory instances of awe, with the ultimate goal of gaining a complete picture of the cerebral building blocks of this complex emotional experience. This ambitious aim is reached via the development of a new multimodal research protocol that integrates immersive nature-based VR scenarios, clinical questionnaires, EEG, and TMS.

In the proposed protocol, to provide evidence of how neuronal information propagates within the brain circuitry during awe experiences, participants perform an EEG study while navigating in natural VR environments validated to elicit awe (32). To monitor awe effects on the excitability of brain networks, after each VR experience, a TMS-EEG study is performed by sending TMS pulses to the left dorsolateral prefrontal cortex (DLPFC), which is believed to be affected by awe (25). Participants are then asked to fill out questionnaires addressing their emotions. We hypothesize that this innovative multimodal approach can provide key evidence of the psychological and neural correlates of awe experiences, which paves the way to plan experiential interventions for the promotion of mental well-being.

## Methods

### Study aims

The SUBRAIN study was born out of a multidisciplinary collaboration between biomedical engineers, psychologists, psychiatrists, and philosophers. It aims to provide unprecedented knowledge on the physiological emotional and neural mechanisms underlying the awe emotion. The following specific aims are pursued: (i) to test the feasibility of a multimodal neuro-centered experimental approach for studying the neurobiological bases of the awe experience in a laboratory environment, (ii) to explore brain responses to different VR experiences of awe, and (iii) to identify brain mechanisms linked with type and intensity of awe. The research protocol has been registered with Open Science Framework (https://osf.io/m47du).

### Study population

Fifty young adults from the general Italian population will take part in the study. Recruitment started on February 18^th^, 2022, and is currently ongoing. The sample recruitment is performed via word of mouth. Inclusion criteria are (i) 20-40 years of age, (ii) normal or corrected vision, and (iii) capability to give informed consent and to comply with the study procedures. Exclusion criteria are (i) balance or vestibular disorders, (ii) severe migraine/headache, (iii) pregnancy, (iv) metal/electric implants in the head-neck district, (v) severe cognitive deficits, (vi) medical conditions predisposing to greater epileptic risk or side effects during TMS (including personal or family history of epilepsy, brain ischemic events, neurological diseases, neurosurgical interventions, vascular or orthopedic head-neck surgery, major head trauma, severe migraine or headaches), (vii) heart, respiratory, kidney, liver failures or immuno-suppression, (viii) substance use disorders, (ix) history of psychotic or mood disorders based on DSM-5, and (x) intellectual disability. Participation in the study is conditional on the signature of the written informed consent to the study protocol and fulfillment of the eligibility criteria.

### Study design

This is an Italian, single-center, one-harm, non-randomized interventional study. The study procedure involves four main phases, (i) screening and enrolment, (ii) pre-experimental assessment, (iii) VR experimental assessment, and (iv) post-experimental debriefing, which are schematized in Fig.1 and detailed in the homonymous subsections of “Interventions”.

**Fig. 1.**
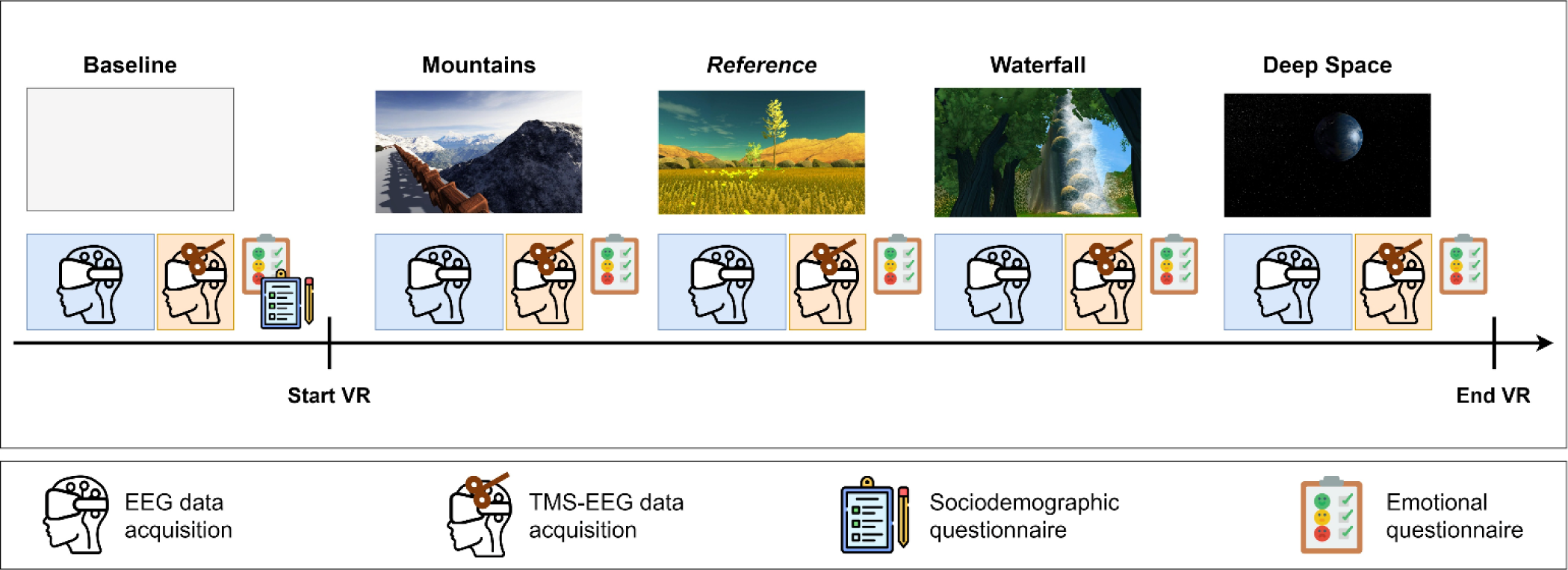
Experimental protocol. EEG and TMS-EEG data are recorded at baseline, along with sociodemographic and emotional questionnaires. EEG data is recorded during each VR scenario, and TMS-EEG is recorded right after the end of the navigation. Afterward, emotional questionnaires are administered.

The study protocol conforms to the Helsinki Declaration and has been approved January 26^th^, 2021, by the competent Ethical Committee of the Fondazione IRCCS Ca’ Granda Ospedale Maggiore Policlinico, Milan (OSMAMI-26/01/2021-0002688-U).

### Study material

#### Questionnaires

1. Case Report Form (CRF): ad-hoc questionnaire for collection of biographic, socio-demographic, and anamnestic information, administered to aspirant participants to check their fulfillment of the inclusion criteria and to investigate any previous experiences with VR environments.
2. short Positive And Negative Affect Scale (PANAS): a brief version of the homonymous scale that measures two main clusters of affective experiences, positive affect and negative affect, by rating 20 adjectives on a 5-point Likert scale (33).
3. Single Item Emotion (SIE) scale: a questionnaire that rates eight emotions, including awe, on a 7-point Likert scale (22).
4. Short Awe Scale (SAS): brief 7-item version of the Awe-Experience Scale (34), which explores the intensity of perceived awe (35).
5. Hamilton Depression Rating Scale (HDRS): a 17-item measure of depression severity rated by the clinician (36).
6. Visual Analog Mood Scale (VAS): a single-item measure of feeling state rated by the participant (37).

### VR equipment and scenarios

The VR system is composed of an Oculus Rift DK2 (Oculus VR LLC, Irvine, CA, United States) equipped with headphones and a Microsoft Xbox controller and connected to a computer with an NVIDIA GTX1070 graphic card and Intel i7 GPU. The Oculus Rift DK2 is a head-mounted display with a 1920×1080-pixel resolution and 90 Hz frame rate. Four interactive and immersive VR scenarios modeled using the Unity software (version 5.5.1) are employed. All scenarios represent natural scenes; three of them have been validated to induce different instances of awe, whereas the remaining one is a neutral, reference condition (22). The three awe-inducing scenarios represent natural scenes of (i) a forest with tall trees, (ii) high mountains, and (iii) the earth viewed from deep space. The neutral reference scenario represents a green grass with few flowers and trees. In the (i) and (ii) scenarios, environmental sounds consistent with the virtual landscapes are added to the visual stimuli with an intensity proportional to the participant’s proximity to the sound source.

### TMS-EEG equipment

The equipment is composed of a Geodesic EEG system 400 (GES 400, Electrical Geodesic Inc., Philips, the Netherlands) equipped with a TMS-compatible 64-channel Microcel Geodesic sensor net (GSN 100, Electrical Geodesic Inc., Philips, the Netherlands). The EEG device is combined with an STM9000 TMS system (Ates Medica Device s.r.l, Italy) equipped with a 70-mm butterfly cooled coil and with the NetBrain 9000 neuronavigation system (EBNeuro S.p.A, Italy).

### Interventions

The four phases of the study procedure are described in the following dedicated sections.

### Screening and enrolment

In this phase, the aspirant participant is informed of the study procedures and freely decides whether to consent to participate in the study. If so, after the signature of the written informed consent, a univocal code is assigned to the aspirant participant. Eligibility criteria and previous experiences with VR are investigated via the administration of the CRF questionnaire. If inclusion criteria are met, the participant is invited to a second visit that includes the three remaining phases.

### Pre-experimental assessment

The participant’s emotional status is retrieved via the short PANAS, SIE, and SAS questionnaires. The participant’s mood is explored via the HDRS and VAS questionnaires. The experimental setup is prepared through the montage and functional check of the VR, TMS, and EEG devices. Eyes-closed resting-state EEG recordings (lasting 2 minutes) and TMS-EEG recordings (lasting about 3.5 minutes) are performed. More details can be found in the “TMS-EEG protocol” section.

### VR experimental assessment

After receiving instructions about the VR experiment, the participant navigates the VR scenarios (three awe-inducing and one of reference) in a counterbalanced order, for about 3 minutes each. Before each experience, the participant is instructed on how to navigate and is asked to keep her/his eyes closed until the scenario has started. EEG signals are continuously recorded during navigation in each VR scenario. An event on the signal is marked at the moment in which the participant opens the eyes and starts the navigation. In the awe-inducing scenarios, a further event is marked when the maximum awe intensity is reached, according to the psychologist, after which the navigation continues for at least 30 seconds. Right after, the participant is requested to stay still, looking at the VR-induced landscape in front of her/him, while TMS-EEG recordings are performed (lasting about 3.5 minutes, see “TMS-EEG protocol”). After the TMS-EEG session, the participant’s emotional and mood status is assessed through the re-administration of the short PANAS, SIE, SAS, and VAS scales.

### Post-experimental debriefing

At the end of the VR experiment, main participants’ perceptions and feelings concerning the VR experiences are captured through a debriefing with the psychologist, which is recorded upon the participants’ consent.

### TMS-EEG protocol

During both EEG and TMS-EEG sessions, EEG signals are recorded at a sampling frequency of 1000 Hz. During the TMS-EEG sessions, EEG signals are continuously recorded while 105 TMS pulses are delivered over the left DLPFC at 0.5 Hz and with a stimulus intensity around 120% of the individual resting motor threshold (rMT). In the pre-experimental phase, the rMT is measured as the minimum intensity needed to obtain 5 out of 10 consecutive right-index movements when stimulated by the left primary motor cortex. The stimulation site of the DLPFC is then defined as an electrode-free point between electrodes E12 (F3) and E15 (FC3), 5 cm ahead of the stimulation motor point. The coil is placed tangentially to the scalp and perpendicular to the EEG wires’ direction in order to reduce the TMS-induced decay artifact (38,39). To ensure that the stimulation point is inside the DLPFC and to increase the intra-subject reproducibility of the stimulation across different TMS-EEG sessions, the neuronavigation system is used. Through preliminary tests, the stimulation point location and intensity are chosen considering the TMS-related artifacts on the EEG, the comfort of the patient, and using as anatomical reference the middle frontal gyrus defined using the Atlas69 atlas on a template image of the neuronavigator. According to the listed parameters, the stimulation point location and coil orientation are saved in the neuronavigator and replicated in the following TMS-EEG sessions. Lastly, to reduce the auditory evoked potential evoked by the TMS pulses, a white noise signal is reproduced during the TMS-EEG sessions through the headphones of the Oculus Rift DK2. The volume of the white noise is adjusted for every participant to cover the TMS clicking noise and accommodate their comfort.

### Sample size calculation

The SUBRAIN study is purely explorative, therefore there is a lack of hypotheses on the size of the physiological behavioral and brain responses to the innovative VR protocol. For these reasons, the sample size calculation is based on the financial resources available for the study conduction. The outcome of the proposed experimental protocol will be measured in terms of the possibility of (i) effectively sending the TMS pulses to the target region, taking into account the VR equipment, (ii) obtaining good quality EEG signal during the VR protocol to extract features of interest, and (iii) effectively removing TMS-induced artifacts from the EEG signals. Considering the ISO guidelines of the technical committee ISO/TC 210 (“Quality management and corresponding general aspects for medical devices”) regarding usability testing of medical devices, we anticipate that the use of the new experimental protocol on 5/10 subjects will allow most (> 70%) of the technical problems of integration of the three systems (VR, EEG, and TMS) to emerge. These problems will be examined and addressed to refine the experimental protocol, which will be applied to the rest of the study sample. Finally, the study’s statistical power, in terms of emotional-behavioral and cerebral correlates of experiences in VR, will be examined and discussed retrospectively.

### Data management plan

Experimenters guarantee the anonymity of the volunteers, treating their common and sensitive data, excluding the personal ones, and avoiding their association with the person by using a pseudonymized form. At the beginning of the experiment, the experimenters associate a code that identifies the subject’s data during all the experimentation phases. The experimental data, including signals, socio-demographical and anamnestic data, and questionnaires, are recorded, analyzed, and preserved along with the code. The file associating participants’ codes and their identification data and the one containing experimental data associated with the participants’ codes are stored on a password-protected computer. Only experimenters and authorized personnel can link the code to the name of the volunteer. Data is recorded, processed, and stored by electronic and non-electronic devices and will be disseminated only in strictly pseudonymized form, such as through scientific publications and conferences.

### Primary endpoints

Development of the experimental and innovative setup based on the successful combination of VR, TMS, EEG, and clinical questionnaires to induce different instances of the awe experience and study its emotional-behavioral and cerebral correlates. Particularly, (i) setup, test, and refinement in a pilot sample (n=6), and usage in an independent sample (n=44), and (ii) identification of emotional-behavioral and brain functional mechanisms, including neuronal oscillatory activity, cortical excitability, and pathways of propagation of neuronal information, underlying the awe experience.

### Secondary endpoints

Identification of specific emotional-behavioral and brain functional mechanisms underlying the different VR-induced awe experiences. Specifically, investigation of (i) the different types and intensities of awe inspired by VR scenarios, (ii) the neural correlates of the subjective awe experience, reported in the questionnaires, independently of the VR scenario itself, and (iii) the specific neural correlates of each VR scenario, by means of comparison between the sublime-inducing VR scenario and the reference one.

### Safety considerations

The project does not involve administering drugs or other substances or invasive clinical practices. Therefore, no adverse events are expected. The interventional design of the study is exclusively due to the use of the innovative VR-TMS-EEG experimental setup, whose components are non-invasive. TMS stimuli, when administered repetitively, several times per second, can induce seizures in predisposed subjects (0.003% of cases). Such seizures are of short duration and leave no consequences. The protocol involves sending TMS stimuli at intervals of more than one second apart. In addition, personal or family history of epilepsy, neurological diseases, neurosurgical interventions, and major head trauma that increase epileptic risk constitute exclusion criteria for the study. At stimulation frequencies such as those used in our study, the risk of seizure induction in subjects not predisposed to develop seizures is negligible (less than 2 cases per 1000 stimulation sessions). The prolonged use of VR can produce discomfort, such as nausea, dizziness, and increased muscle fatigue.

### Statistical analyses

EEG and TMS-EEG data pre-processing is carried out using BrainVision Analyzer (BrainProducts, Germany) and EEGLAB, an open-source toolbox running in Matlab R2022a (Mathworks, Inc., Natick, Massachusetts, USA). Neuronal and emotional questionnaire data are analyzed using in-house Matlab scripts.

Statistical data analyses aim to evaluate the differences, in terms of emotional questionnaire scores and neural correlates, induced by (i) the experience of awe, comparing the awe-inducing VR scenarios with the reference VR one and (ii) the VR, comparing the reference VR scenario with the baseline. A further objective is to correlate the neuronal changes with the emotional ones. To achieve these goals, (i) the values of intensity of emotional dimensions recorded with the SAS, SIE, and PANAS questionnaires are compared using paired t-tests or Friedman statistics across conditions to evaluate the subjective effect of the different VR scenarios. Brain complexity, power, and functional and effective connectivity features are extracted from the EEG signal and used to evaluate differences of multiple cerebral dimensions among the experimental conditions, ultimately providing a wide comprehension of the neural bases of the awe emotion. Amplitude and latency of TMS-evoked potentials (TEPs) of interest (40) and TMS-evoked effective connectivity indices (41) are calculated from the TMS-EEG data and compared among the experimental conditions. Power and connectivity analyses are performed in the main EEG frequency bands (delta: 1-4 Hz, theta: 4–8 Hz, alpha: 8–13 Hz, beta: 13–30 Hz, and gamma: 30–50 Hz). Multi-scale connectivity features are extracted using graph theory. Finally, the association between EEG and TMS-EEG features and scores in the emotional questionnaires is further investigated via generalized linear models.

### Current status

The setup, test, and refinement of the experimental protocol, which is a primary endpoint of the SUBRAIN study, have been achieved by successfully combining TMS, EEG, VR and by refining their integrated usage in the experimental session. Up to now data from ten participants (5M, 27.2 ± 3.4 years; 5F, 28.4 ± 5.2 years), including six participants for the pilot study, have been recorded. Using the proposed protocol, a pilot study on six healthy controls showed the possibility of extracting standard TEPs, in line with the literature, when stimulating the left DLPFC at baseline (40). This pilot study has been followed by the inclusion of the neuronavigator in the experimental equipment, which has allowed us to improve the intra-subject reproducibility of TMS pulses sent after the navigation in different VR scenarios. Also, to ensure the robustness of the analyses, the nauseous state of the subject, found especially after the reference scenario, was tracked with annotations with the goal of incorporating it into statistical models. The endpoints relative to the investigation of the psychological and brain correlates of the VR-induced awe experience are still under study. A preliminary analysis showed a correlation between the reported intensity of awe emotion and the amplitude or latency of specific TEPs of interest. Also, different VR scenarios reported different brain responses regarding both EEG power spectrum and EEG connectivity. Specifically, the preliminary findings suggest an increase of connectivity in delta and theta bands for space and mountain scenarios.

### Data availability

Socio-demographic data, psychopathological and emotional questionnaire scores, EEG, and TMS-EEG data recorded with the described protocol are protected by privacy and will be available from the corresponding author upon reasonable request, after the signature of a formal data sharing agreement, in an anonymous form.

## Discussion

Through the incorporation of pioneering quantitative, multidisciplinary, and multimodal experimental and analytical methodology, this protocol aims to enhance the currently limited understanding of the cerebral foundations of the awe emotion. We seek to present fresh insights into the collaboration of various brain regions in generating the profound, enigmatic, and transformative experience of awe. This protocol employs a meticulously designed framework that integrates VR with neural techniques, including EEG and TMS, alongside emotional questionnaires, to investigate the mechanisms underpinning VR-induced awe experiences.

The preliminary data obtained so far demonstrates the proposed protocol’s feasibility and reproducibility, unveiling distinctions of neural mechanisms across various VR-induced awe scenarios. The use of interactive and immersive VR as an awe-inducing medium enables the participant’s full and ecological immersion in the experience, as previously demonstrated (20,22), whereas the combination of EEG and TMS enables the extensive exploration of the cortical changes induced by diverse instances of awe through VR scenarios, offering novel insights into this intricate emotional experience. Integrating different neural techniques and tailored psychological questionnaires contributes to unveiling the specific emotion under study and opens the door to new insights into the brain mechanisms underlying the perception of complex emotions.

A central question arising from this investigation pertains to the potential use of experiential interventions based on the Hry elicitation of awe emotion to promote mental health. At a biological level, awe appears to decrease default mode network activity (24), sympathetic activation (21), and pro-inflammatory cytokines (42). Moreover, awe seems to reduce the ruminative and self-referential thinking characterizing some psychiatric disorders, such as Major Depressive Disorder (MDD). The natural extension of the proposed study will employ this experimental framework in the study of MDD to gain a deeper understanding of the neurobiological correlates of awe in affected individuals, investigating the therapeutic potentiality of awe-inducing VR scenarios.

While focusing on the awe emotion, our approach offers a reproducible framework adaptable to diverse research domains, especially those exploring immersive experiences. However, several limitations merit consideration. VR could induce sickness and nausea in some patients (43) which could, in turn, impact the experience, altering the experience of awe and its beneficial effects. A questionnaire evaluating the level of nausea after each VR scenario should be implemented to keep track of this phenomenon. Our study implemented three different awe-inducing scenarios designed to enhance the vastness dimension (23,44), however, other awe dimensions could be investigated, increasing the number of possible new VR scenarios. Finally, an intrinsic limitation is posed by the use of EEG to investigate brain activity, due to its inability to unambiguously detect the activity of deep neural sources, such as EEG signal changes generated by areas of the DMN. Nevertheless, the possibility to combine EEG with VR and TMS makes this technique the most suitable for our research purpose, that is, to understand the functional brain correlates of the awe emotion elicited in an immersive environment.

## Conclusion

Our study results from an unprecedented synergistic integration of complementary expertise in bioengineering, psychology, philosophy, and psychiatry. SUBRAIN adopts an innovative, quantitative, and multimodal experimental and analytical approach to increase the currently limited knowledge of the cerebral bases of the awe experience. With this protocol, we intend to provide new evidence on how multiple brain areas can work together to produce the powerful, mysterious, transformative awe emotion.

## Acknowledgments

We wish to thank Viviana Pescuma, Yara Massalha, and Andrea Anastasi for the crucial contribution to the data acquisition phase.

